# A Randomized, Double-blind, Placebo-controlled Crossover Pilot Study of the Effect of Metformin on Airway Glucose in COPD: The Metformin and Airway Glucose In COPD (MAGIC) Trial

**DOI:** 10.1101/2024.11.13.24317228

**Authors:** Eva Fiorenzo, John S. Tregoning, Isaac Fahidi, Michael R. Edwards, Tata Kebadze, Patrick Mallia, Sebastian L. Johnston, Hugo A. Farne

**Affiliations:** National Heart and Lung Institute, Imperial College London; Department of Infectious Disease, Imperial College London

**Keywords:** Chronic Obstructive Pulmonary Disease, COPD, Exacerbations, Metformin, Glucose, Randomized Controlled Trial

## Abstract

**Background and Objective:** Patients with COPD have elevated levels of airway glucose. This may increase the risk of bacterial infection leading to exacerbation and disease progression. We aimed to test whether treating COPD patients with metformin, an oral hypoglycaemic used in diabetes, reduced airway glucose.

**Methods:** In this randomized, double-blind, placebo-controlled crossover pilot study, we assigned patients with COPD to two 3-month periods of treatment with 1g metformin or placebo twice daily, followed by the alternate treatment after a 2-week washout period. Patients were required to be free of infection, antibiotic or oral steroid treatment in the 8 weeks prior to enrolment. The following were collected at baseline and monthly thereafter: sputum (lower airways sample), nasal/upper airway lining fluid samples using synthetic absorptive matrix (SAM), spirometry, and St Georges Respiratory Questionnaire (SGRQ) and COPD Assessment Test (CAT) scores. The primary outcome was a change in sputum glucose following 3 months treatment with metformin compared to 3 months of placebo.

**Results:** 14 patients were randomised of whom 3 completed the study, mainly due to withdrawals for exacerbations (8/11 withdrawals). In the n=4 patients who completed 3 months metformin treatment, there were no significant changes in sputum or nasal glucose compared to baseline. Metformin did not affect COPD symptom scores or spirometry.

**Conclusions:** Treatment with metformin in this underpowered pilot study did not significantly reduce airway glucose in COPD. Larger studies are required to definitively evaluate this.

**SUMMARY AT A GLANCE:** Elevated airway glucose is associated with bacterial infection in chronic obstructive pulmonary disease (COPD) exacerbations. We tested whether metformin, a drug used to treat diabetes, reduced airway glucose in patients with COPD. Recruitment/retention difficulties led to an underpowered study, which observed no difference between metformin and placebo on airway glucose.

## INTRODUCTION

Chronic obstructive pulmonary disease (COPD) affects an estimated 400 million people globally.^(1)^ As well as chronic respiratory symptoms, patients with COPD suffer from episodes of acutely increased symptoms called exacerbations. These can be triggered by viral infections, bacterial infections and pollution, with bacteria detected in 50-60% of exacerbations. Bacterial infection of the lower airways is also associated with airway inflammation, disease progression and mortality.^(2)^ Exacerbations with bacterial infection may require antibiotics, yet with antibiotic resistance a growing problem, there is a need for alternative treatments. One therapeutic target is airway glucose; increased glucose levels have been observed in the nasal lavage and sputum of stable COPD patients with further increases during viral/bacterial COPD exacerbations.^(3)^ In primary human airway epithelia models, lower levels of airway glucose impaired *Pseudomonas aeruginosa* growth in vitro.^(4)^ Furthermore, in mice, hyperglycaemia promoted *P. aeruginosa* and *Staphylococcus aureus* growth in the airways.(^4, 5)^ This suggests that levels of airway glucose affect microorganism growth and survival. It may also explain why human bronchial epithelial cells increase glucose uptake in the presence of cytokines or pro-inflammatory mediators; the subsequently reduced airway glucose levels may prevent infection.^(6)^

A potential approach to modulating airway glucose is with existing oral hypoglycaemic drugs, developed and licensed for the treatment of diabetes. Metformin is one such drug and has been safely used in patients with COPD and diabetes without causing hypoglycaemia.^(7)^ In patients with co-morbid COPD and diabetes, metformin treatment is associated with a lower risk of COPD-related hospitalisation and death, particularly after 2 years use, and more so than other hypoglycaemic agents including insulin.^(8-11)^ A 24% reduced risk of COPD exacerbation has been reported following up to 180 days metformin use^(12)^ as well as a 28% and 37% reduced risk of inpatient visits with COPD after 91-180 days or 181-365 days metformin use respectively. ^(13)^

The mechanism by which metformin reduces COPD exacerbations remains unclear. One study found that 6 months of metformin treatment did not improve spirometry or alter exhaled nitric oxide, despite being associated with significant changes in symptom scores.^(14)^ Furthermore, few studies have examined the effects of metformin in COPD patients without diabetes. 1 month of treatment with metformin in patients with COPD admitted for an exacerbation who did not have diabetes, had no impact on blood glucose (which was modestly raised in both the metformin and placebo-treated groups), blood levels of the inflammatory marker C-reactive protein, or scores on the COPD Assessment Test (CAT) or Exacerbations of Chronic Pulmonary Disease Tool (EXACT).^(15)^ However treatment began at hospital presentation, some days after viral infection and symptom onset, and blood but not airway glucose levels were measured. We therefore undertook a pilot trial to determine whether metformin altered airway glucose in COPD and to assess the feasibility of conducting a full-scale trial of the efficacy of metformin prophylaxis in reducing COPD exacerbations.

## METHODS

### Human/Animal Ethics Approval Declaration

IRAS Number: 247421

Westminster Research Ethics Committee reference: 19/LO/0633

### Study Design

The Metformin and Airway Glucose In COPD (MAGIC) trial was a double-blinded, randomised, placebo-controlled cross-over trial in COPD. Recruitment began 17 May 2022 and ended 01 June 2023. Participants were randomised to metformin or placebo for 3 months, followed by a 2-week washout period before crossing over treatment arms. The participants attended a baseline visit followed by monthly visits until the third month of treatment was completed. This was repeated after crossing over. At these visits, symptoms, quality of life (QOL) and spirometry were assessed, and sputum and nasal synthetic absorptive matrix (SAMs) samples collected.

### Participants

Patients aged 40-85 years with a clinical diagnosis of COPD confirmed on spirometry (post-bronchodilator FEV_1_/FVC <70%) and a smoking history of at least 15 pack years were eligible for the study. Exclusion criteria included a diagnosis of diabetes (previously or on screening bloods), treatment with metformin, contraindications to metformin (e.g. previous allergic reaction to metformin, body mass index <18.5kg/m^2^), and a history of infection, antibiotic and/or oral corticosteroid use in the previous 8 weeks. See supplementary material for full inclusion and exclusion criteria. Written consent was obtained prior to participation.

### Intervention, Randomisation and Masking

The study drugs were supplied by Sharp Clinical Services (Powys, UK), who produced visually identical metformin and placebo tablets with the same packaging, differentiated only by a unique pack ID. Eligible subjects were randomly assigned to either metformin or placebo twice daily in a 1:1 ratio, according to a sequential randomisation list generated by a statistician working independently of the trial that was encoded in the study database and concealed up to allocation. Subjects and study investigators were blinded to treatment allocation from randomisation until database lock. Treatment identity was concealed by using placebo identical in packaging, labelling, schedule of administration, appearance, taste and odour to metformin. Metformin was dosed at 500mg twice daily for one week to check tolerability, increasing to 1g twice daily thereafter for the remainder of the 3 months. Patients were considered compliant if ≥80% of investigational medicinal product (IMP) was taken.

### Outcomes

The primary outcome was change in sputum glucose following 3 months’ treatment with metformin. Glucose was measured from sputum plugs using a commercial assay, the Amplex Red Glucose Assay Kit (Thermo Fisher Scientific, UK). Secondary outcomes included glucose concentrations in nasal SAM strips, COPD Assessment Test (CAT) and St George’s Respiratory Questionnaire (SGRQ) scores, and forced expiratory volume in 1 second (FEV_1_).

### Statistical Analysis

A sample size calculation indicated that 30 subjects were required to detect a reduction of 30% in airway glucose, based on the observation of similar reductions in airway glucose following metformin treatment in mice,^(16)^ at 90% power using a two-sided test at the 5% significance level. This was grossed up to 40 subjects to account for withdrawals.

Data were analysed using GraphPad Prism 9 (GraphPad Software, USA). Continuous variables are presented as medians and interquartile ranges, with the Mann-Whitney U test used to analyse between groups and Wilcoxon signed-rank test for paired analyses. Differences were considered statistically significant at p<0.05. All p values are two sided.

## RESULTS

Between 17 May 2022 and 01 June 2023, 14 participants were enrolled. 11 were subsequently withdrawn spread evenly across the metformin and placebo treatment arms, 8 due to lower respiratory tract infections (Figure 1A). 3 subjects completed 3 months of both metformin and placebo; 8 volunteers completed 3 months of a single arm (4 per arm; Figure 1B). In total, there were 16 sets of samples following 1 month’s treatment of either placebo or metformin (8 and 8 respectively); 12 from subjects who completed 1 month in the study and a further 4 from subjects who crossed over and completed 1 month in the other treatment arm). The trial was ended following expiry of the study drug on 31 May 2023.

**Figure 1.**
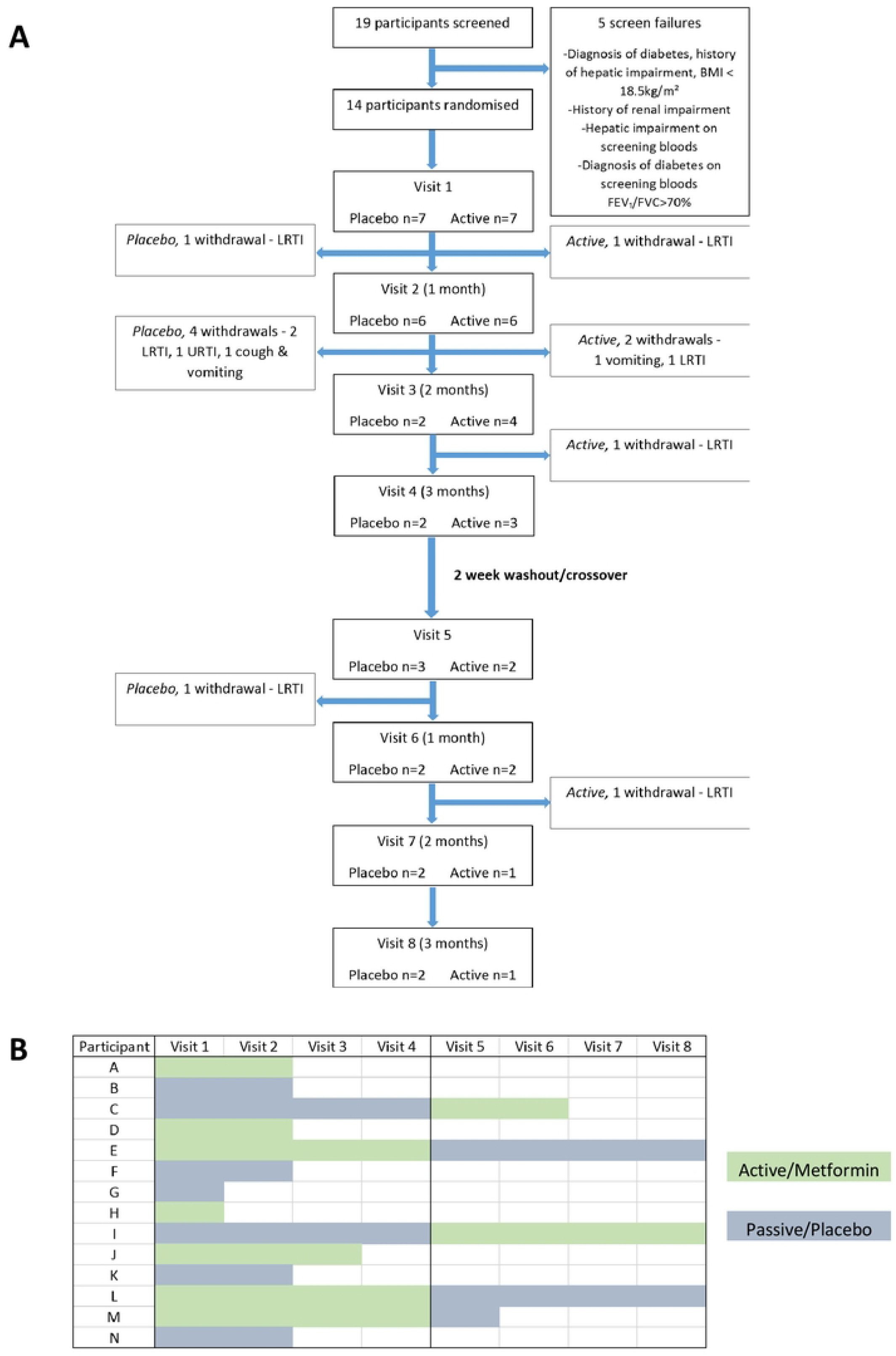
Outline of participation in MAGIC study. Consort diagram (A). Timelines of individual participants of study (B).

There was no significant change in the primary outcome, sputum glucose following 3 months treatment with metformin (median sputum glucose 68.92 (IQR 16.3 to 131.1) μM, either compared to baseline 5.3 (IQR 0.02 to 18) μM (p=0.13; n=3) or 3 months placebo (median sputum glucose 12.2 (IQR 10 to 13) μM. Furthermore, there was no change in sputum or nasal glucose with metformin treatment at any timepoint from baseline to 3 months (Figures 2A-B), including when only analysing patients who completed 3 months of either arm of treatment (Figures 2C-D). We then analysed the change in sputum and nasal glucose after 1 month’s treatment compared to baseline and found no significant change (Figures 2E-F).

**Figure 2.**
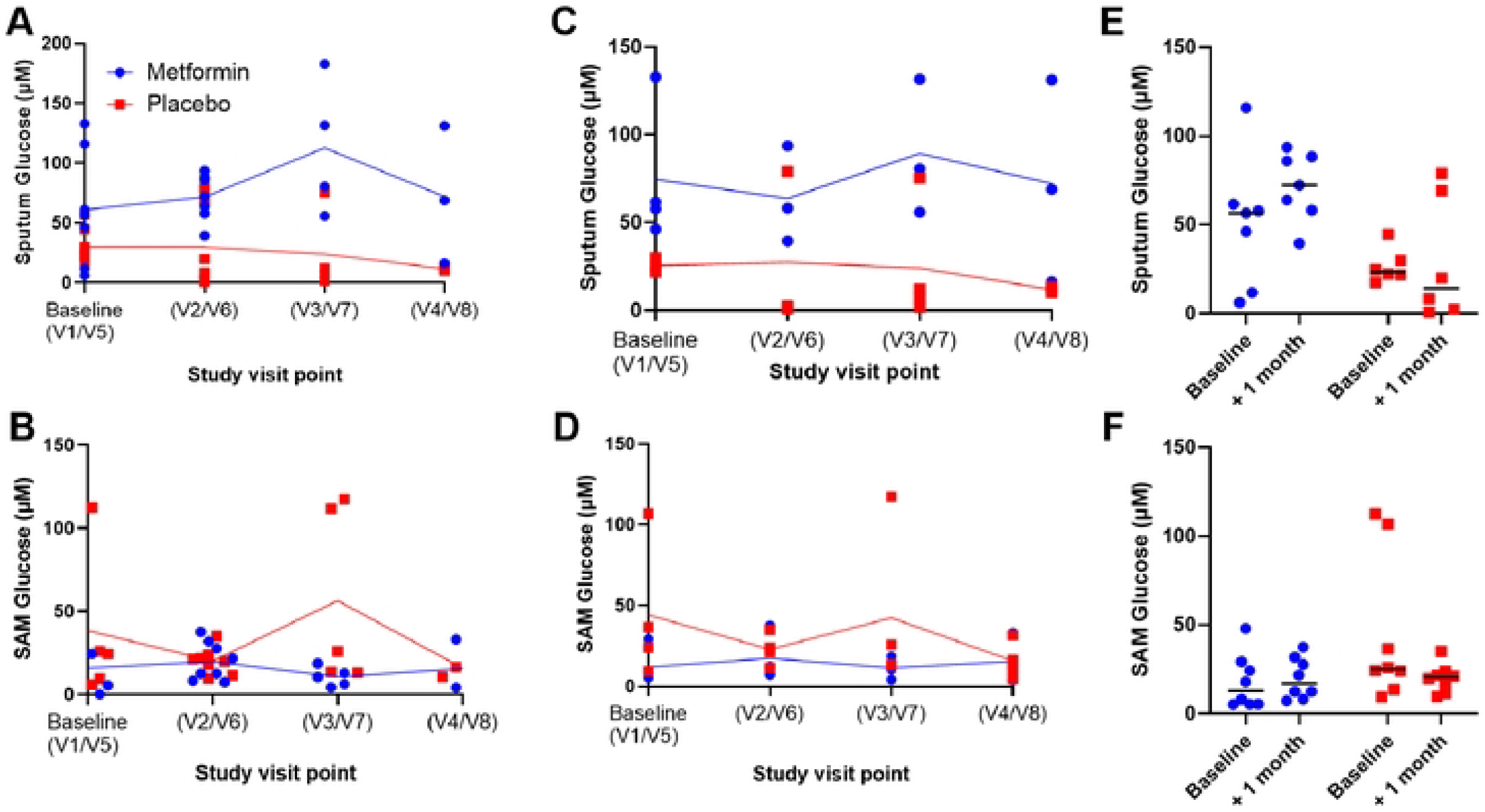
Effect of metformin on glucose in upper and lower airways. Volunteers with COPD were assigned to either active or placebo with a washout period before switching. Glucose levels were measured in sputum (A, C, E) samples and synthetic absorptive matrices (SAM) collected from the nose (B, D, F). Glucose levels over the time course of the study for all participants measured (A, B); levels in individuals who complete a full 3-month participation (C, D). Comparison of glucose levels at baseline (V1/V5) and one month later (V2/ V6). At baseline N=9 participants in active, N=10 in placebo, n=4 participants finished a 3-month course of either metformin or placebo. Symbols represent individual participants and lines medians, statistical analysis was performed by Mann-Whitney test (A-D) and Wilcoxon signed-rank test (E, F).

We additionally evaluated whether metformin affected airway function or symptoms of COPD, assessed by the CAT and SGRQ questionnaires. There were no significant differences in any measurement at baseline between the two arms (Figure 3A), and no difference after 1 month of treatment compared to baseline or between placebo and metformin treatment (Figures 3B-F).

**Figure 3.**
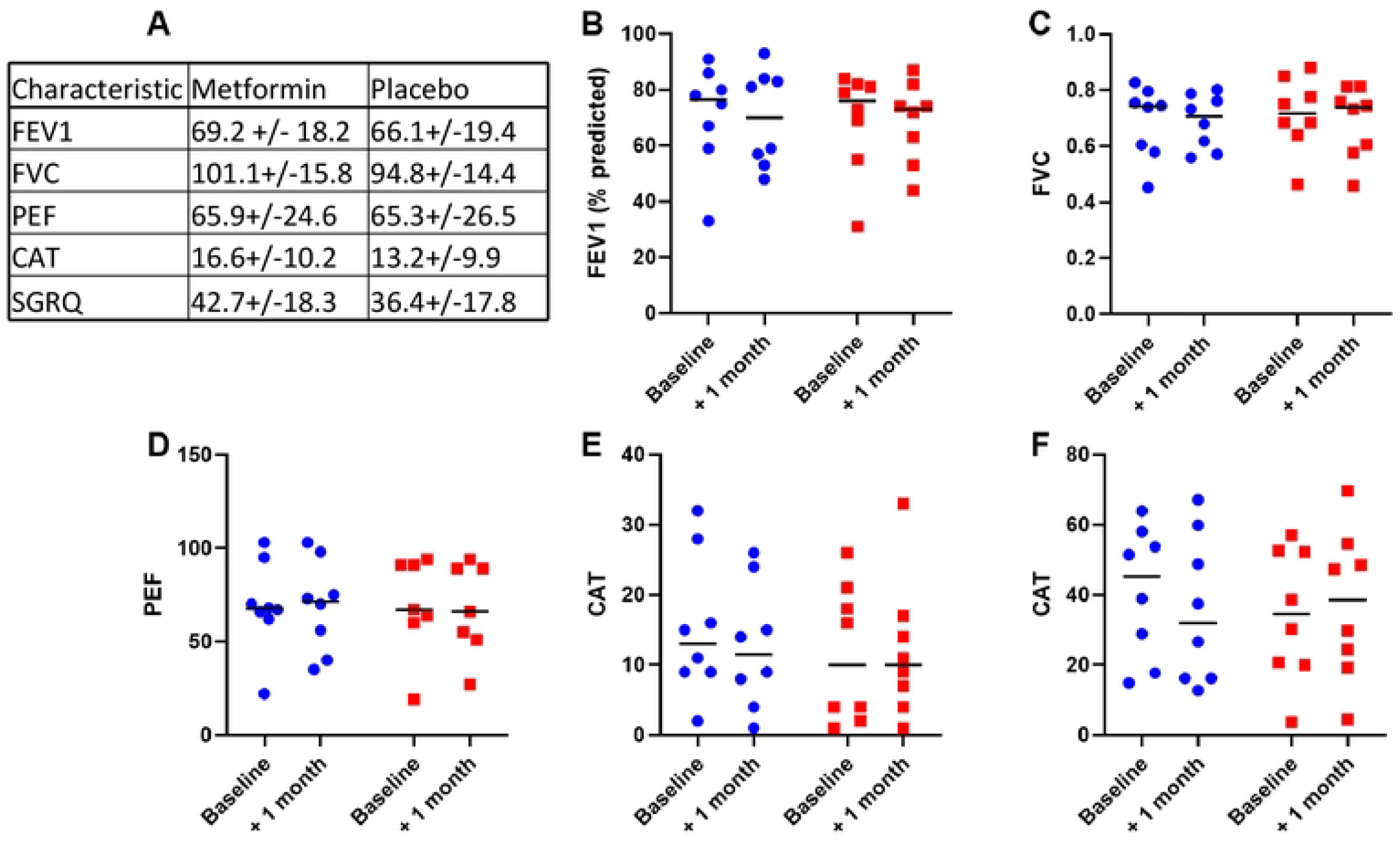
Metformin had no impact on measures of airway function at one month. Volunteers with COPD were assigned to either active or placebo with a washout period before switching. Measurements of airway function and symptoms were taken at visit 1 (A). Airway function and symptoms were compared between initial visit 1 (V1) and Visit 2 (V2) of each arm for FEV1 (B), FVC (C), PEF (D), CAT (E) and SGRQ (F). n ≥ 9. Symbols represent individual participants and lines medians, statistical analysis was performed by Wilcoxon signed-rank test.

## DISCUSSION

This pilot study adds to the literature on metformin in COPD by showing for the first time that up to 3 months of metformin treatment does not affect glucose levels in upper (nasal) or lower (sputum) airways samples, albeit in small numbers of patients. If born out in larger prospective studies, this would suggest that metformin does not reduce COPD exacerbations by modulating airway glucose. It may be that metformin does not reduce exacerbations at all; the reduced rate of COPD exacerbations seen in patients with comorbid diabetes on metformin ^(8-11)^ is currently an association and not known to be causal.

Furthermore, a comparison of airway glucose in patients with only COPD not on metformin against those with COPD and type II diabetes on metformin has not been made. Like other studies, we failed to see any impact of metformin on FEV_1_ or FVC,(^14, 17^) even though high airway glucose levels are associated with airway hyperresponsiveness.^(18)^

The main limitation of our study is the small sample size, due to a combination of under-recruitment and a high number of withdrawals. Study recruitment was delayed due to the COVID-19 pandemic and limited by the expiry date of the IMP when the study resumed, hence only 14 out of a target 40 patients were recruited. We subsequently had a high drop-out rate owing primarily to the exclusion of patients who developed a lower respiratory tract infection (8 of the 11 withdrawals). These were excluded on the basis that both lower respiratory tract infections and the oral steroids often used to treat them (alongside antibiotics) are known to alter airways glucose and could therefore confound the results. The inevitable consequence, however, in a population of patients who suffer frequent exacerbations (particularly in the autumn and winter when most of the subjects were enrolled) was a high number of withdrawals. Future studies could consider including these participants and performing subgroup analyses on samples from non-infective timepoints. However, even with our small numbers, there was no trend towards a difference in airway glucose with metformin treatment.

The sputum results may have been affected by the poor quality of samples with low cell counts and high cell death, likely due to immediate freezing of samples after collection. The same would not be applicable to the nasal SAM samples, although these may not reflect sputum (lower airways) glucose as differences have previously been observed in the biochemicals measured in sputum and nasal SAM samples collected at the same time.^(19)^

Outcomes of previous studies have suggested that longer periods of treatments such as 6 months may be required to detect statistically significant changes in patient symptoms.^(14)^ At a minimum, 91 days of oral anti-hyperglycaemic treatment has been required to reduce the risk of exacerbation, although this will be affected by the frequency of exacerbations.^(13)^ However exacerbations are relatively infrequent events and for this reason we chose a mechanistic outcome, airway glucose, that should change sooner. Nonetheless future research may want to investigate levels of airway glucose in patients with COPD and comorbid diabetes treated with metformin for various durations, to those with COPD but without diabetes. Similarly, a trial of an oral hypoglycaemic intervention might consider studying a longer duration of treatment.

In summary, in this underpowered pilot study we found no effect of up to 3 months metformin treatment on upper or lower airways glucose in COPD. Given the strong epidemiological evidence linking metformin treatment to reduced rates of COPD exacerbation and the growing problem of antibiotic resistance, larger interventional studies of metformin treatment in COPD are warranted, and would ideally include analysis of a possible mechanism (e.g. measurement of airways glucose).

## Data Availability

All relevant data are within the manuscript and its Supporting Information files.

## Author Contributions

EF: Investigation, Writing - Original Draft; IF: Investigation; MRE: Supervision; TK: Investigation; PM: Conceptualization, Funding acquisition; JST: Conceptualization, Funding acquisition, Writing - Original Draft; SLJ: Supervision, Conceptualization, Funding acquisition, Writing - Review & Editing; HF: Supervision, Investigation, Writing - Review & Editing.

## Acknowledgements

NIHR Imperial Biomedical Research Centre (BRC) (P74536)

## Data Availability Statement

Data availability on request.

## REFERENCES

1. Adeloye D, Song P, Zhu Y, Campbell H, Sheikh A, Rudan I, et al. Global, regional, and national prevalence of, and risk factors for, chronic obstructive pulmonary disease (COPD) in 2019: a systematic review and modelling analysis. Lancet Respir Med. 2022;10(5):447–58.

2. Beasley V, Joshi PV, Singanayagam A, Molyneaux PL, Johnston SL, Mallia P. Lung microbiology and exacerbations in COPD. Int J Chron Obstruct Pulmon Dis. 2012;7:555–69.

3. Mallia P, Webber J, Gill SK, Trujillo-Torralbo MB, Calderazzo MA, Finney L, et al. Role of airway glucose in bacterial infections in patients with chronic obstructive pulmonary disease. J Allergy Clin Immunol. 2018;142(3):815–23 e6.

4. Pezzulo AA, Gutierrez J, Duschner KS, McConnell KS, Taft PJ, Ernst SE, et al. Glucose depletion in the airway surface liquid is essential for sterility of the airways. PLoS One. 2011;6(1):e16166.

5. Garnett JP, Baker EH, Naik S, Lindsay JA, Knight GM, Gill S, et al. Metformin reduces airway glucose permeability and hyperglycaemia-induced Staphylococcus aureus load independently of effects on blood glucose. Thorax. 2013;68(9):835–45.

6. Garnett JP, Nguyen TT, Moffatt JD, Pelham ER, Kalsi KK, Baker EH, et al. Proinflammatory mediators disrupt glucose homeostasis in airway surface liquid. J Immunol. 2012;189(1):373–80.

7. Hitchings AW, Archer JR, Srivastava SA, Baker EH. Safety of metformin in patients with chronic obstructive pulmonary disease and type 2 diabetes mellitus. COPD. 2015;12(2):126–31.

8. Ho TW, Huang CT, Tsai YJ, Lien AS, Lai F, Yu CJ. Metformin use mitigates the adverse prognostic effect of diabetes mellitus in chronic obstructive pulmonary disease. Respir Res. 2019;20(1):69.

9. Liang Z, Yang M, Xu C, Zeng R, Dong L. Effects and safety of metformin in patients with concurrent diabetes mellitus and chronic obstructive pulmonary disease: a systematic review and meta-analysis. Endocr Connect. 2022;11(9).

10. Bishwakarma R, Lin Y, Kuo Y, Sharma G. Metformin and Health Care Utilization in Patients With Coexisting COPD and Diabetes. Chest Journal. 2016;150(4).

11. Bishwakarma R, Zhang W, Lin YL, Kuo YF, Cardenas VJ, Sharma G. Metformin use and health care utilization in patients with coexisting chronic obstructive pulmonary disease and diabetes mellitus. Int J Chron Obstruct Pulmon Dis. 2018;13:793–800.

12. P.L. T, Valencia-Hernandez C, Farne H, Bloom C. Effect of metformin on reducing the risk of COPD exacerbations: a UK nested case-control study. Thorax. 2023;78.

13. Wang MT, Lai JH, Huang YL, Kuo FC, Wang YH, Tsai CL, et al. Use of antidiabetic medications and risk of chronic obstructive pulmonary disease exacerbation requiring hospitalization: a disease risk score-matched nested case-control study. Respir Res. 2020;21(1):319.

14. Sexton P, Metcalf P, Kolbe J. Respiratory effects of insulin sensitisation with metformin: a prospective observational study. COPD. 2014;11(2):133–42.

15. Hitchings AW, Lai D, Jones PW, Baker EH, Metformin in CTT. Metformin in severe exacerbations of chronic obstructive pulmonary disease: a randomised controlled trial. Thorax. 2016;71(7):587–93.

16. Gill SK, Hui K, Farne H, Garnett JP, Baines DL, Moore LSP, et al. Increased airway glucose increases airway bacterial load in hyperglycaemia. Sci Rep. 2016;6:27636.

17. Kahnert K, Andreas S, Kellerer C, Lutter JI, Lucke T, Yildirim O, et al. Reduced decline of lung diffusing capacity in COPD patients with diabetes and metformin treatment. Sci Rep. 2022;12(1):1435.

18. Cazzola M, Calzetta L, Rogliani P, Lauro D, Novelli L, Page CP, et al. High glucose enhances responsiveness of human airways smooth muscle via the Rho/ROCK pathway. Am J Respir Cell Mol Biol. 2012;47(4):509–16.

19. Farne H, Groves HT, Gill SK, Stokes I, McCulloch S, Karoly E, et al. Comparative Metabolomic Sampling of Upper and Lower Airways by Four Different Methods to Identify Biochemicals That May Support Bacterial Growth. Front Cell Infect Microbiol. 2018;8:432.

